# Modeling an Integrated HIV Prevention and Care Continuum to Achieve the Ending the HIV Epidemic Goals

**DOI:** 10.1101/2020.03.02.20030254

**Authors:** Samuel M. Jenness, Jordan A. Johnson, Karen W. Hoover, Dawn K. Smith, Kevin P. Delaney

**Affiliations:** Department of Epidemiology, Emory University; Division of HIV/AIDS Prevention, Centers for Disease Control and Prevention

**Author notes:** **Correspondence** Samuel M. Jenness, Emory University, 1520 Clifton Road, Atlanta, GA 30323.

**Keywords:** Mathematical model, Sexual networks, HIV, HIV care continuum, Preexposure prophylaxis, men who have sex with men, Ending the HIV Epidemic

## Abstract

**Objective:** We sought to evaluate which combinations of HIV prevention and care activities would have the greatest impact towards reaching the US Ending the HIV Epidemic (EHE) plan goals of reducing HIV incidence at least 75% by 2025 and 90% by 2030.

**Design:** A stochastic HIV transmission model for men who have sex with men (MSM), calibrated to local surveillance estimates in the Atlanta area, a focal EHE target jurisdiction.

**Methods:** Model scenarios varied HIV screening rates relative to current levels, under different assumptions of how HIV-negative MSM would be linked to PrEP initiation, and also considered improvements to HIV care linkage and retention for those screening positive.

**Results:** A 10-fold relative increase in HIV screening rates (to approximately biannual screening for black and Hispanic MSM and quarterly for white MSM) would lead to 43% of infections averted if integrated with PrEP initiation. Improvements to HIV care retention would avert 41% of infections if retention rates were improved 10-fold. If both screening and retention were jointly improved 10-fold, up to 74% of cumulative infections would be averted. Under this scenario, it would take 4 years to meet the 75% EHE goal and 12 years to meet the 90% goal for MSM in Atlanta.

**Conclusions:** Interventions to improve HIV screening linked with PrEP for those screening negative, and HIV care retention would have a substantial impact on HIV prevention. However, additional interventions may be necessary to reach the EHE goal of a 90% reduction in incidence for Atlanta MSM by 2030.

## INTRODUCTION

Despite the availability of highly effective HIV prevention tools such as preexposure prophylaxis (PrEP) and treatment as prevention (TasP), antiretroviral therapy (ART) coverage in high-risk populations remains low in many areas in the United States [1–4]. The new *Ending the HIV Epidemic* (EHE) plan seeks to reduce HIV incidence nationally by 75% in 5 years (by 2025) and 90% in 10 years (by 2030) by targeting federal resources towards interventions that increase access and uptake of these prevention tools in high-need regions like the Southeast [5]. However, it is unknown what combinations and what amounts of interventions will be needed to meet those goals.

HIV elimination depends on reducing HIV acquisition and transmission rates through achieving adequate levels of ART coverage at the population level [6]. HIV care and prevention continuum frameworks have defined the intermediate steps that lead to goals of viral suppression for those living with HIV, and high levels of PrEP adherence and persistence for those uninfected but at risk [2,7,8]. Mathematical models have then been used to estimate the relationship between continuum levels and future HIV prevention goals [9]. Prior models evaluating the HIV care continuum in the United States have suggested that major improvements to HIV screening, care linkage, and care retention would be needed to achieve substantial reductions in HIV transmission [10–13]. Models focused on HIV-uninfected populations have projected the impact of PrEP given low coverage and poor persistence [14–16]. Despite this strong modeling research base, including recent models that evaluated both HIV prevention and treatment interventions [17], the potential for clinical synergy between the HIV care and prevention continua has not been sufficiently evaluated. Both continua rely on HIV screening, a gateway to PrEP initiation for persons who test negative and ART initiation for persons who test positive [8].

In this study, we developed a mathematical model of HIV with an integrated HIV prevention and care continuum framework in a target population of MSM in the Atlanta area, a focal EHE target jurisdiction. In 2017, Atlanta had the second highest rate of new diagnoses of any metropolitan area in the U.S., and 82% of cases occurred in black and Hispanic MSM [18]. Addressing the joint challenges of high overall HIV incidence and racial disparities is a key priority of EHE implementation activities [19]. Our primary research question was what combinations of increases in HIV screening (alone or as a gateway to PrEP initiation), HIV care linkage, and HIV care retention could meet the EHE goals of 75% and 90% reduction in HIV incidence. These projections may inform HIV implementation science and public health policy about what HIV prevention strategies should be prioritized in high-incidence, high-need settings like Atlanta that are the focus of the new EHE plan.

## METHODS

### Study Design

Our network-based mathematical model of HIV transmission dynamics for US MSM was built with the *EpiModel* platform [20], software for simulating epidemics over dynamic contact networks under the statistical framework of temporal exponential random graph models (TERGMs) [21]. Building on our previous applications to estimate the impact of HIV PrEP for MSM [4,14], current model extensions integrated novel network data streams and expanded the representation of the HIV care continuum. Full methodological details are provided in a Supplemental Technical Appendix [**LINK**].

### HIV Transmission and Progression

This model simulates the dynamics of main, casual, and one-time sexual partners for black, Hispanic, and white/other MSM, aged 15 to 65, in the Atlanta metropolitan area. The starting network size in the model simulations was 10,000 MSM, which could increase or decrease over time based on arrival (sexual debut) and departure (mortality or assumed sexual cessation at age 65). We used primary data from the ARTnet study to fit statistical models for the TERGMs. ARTnet was a web-based egocentric network study conducted in 2017–2019 of MSM in the US, with data from 4,904 respondents reporting on 16,198 partnerships [22]. We included a main effect for geography of residence (city of Atlanta versus all other areas) in models to represent our study target population. Parameters were weighted by census-based race/ethnicity and age distributions to account for sampling biases.

In the TERGMs, predictors of partnership formation included partnership type, degree distributions by partnership types, heterogeneity in degree and assortative mixing by race/ethnicity and age, and mixing by sexual position. Partnership durations were modeled for main and causal partnerships as a set of dissolution rates stratified by partnership type and age group. Other statistical models were fit to ARTnet data to predict frequency of acts within partnerships and the probability of condom use as a function of race/ethnicity, age, diagnosed HIV status, and partnership type.

MSM progressed through HIV disease with HIV viral loads modeled as a continuous attribute. Men could be screened for HIV and initiate ART, which would lower their HIV viral load (VL) and increase their longevity [23,24]. Lower VL with sustained ART use was associated with a reduced rate of HIV transmission probability per act [25,26]. Factors modifying the HIV acquisition probability per act included PrEP use [27], condom use [28], sexual position [29], and circumcision [30].

### HIV Prevention and Care Continuum

We simulated an integrated HIV continuum of antiretroviral-based prevention and care, with HIV screening as the central gateway to either PrEP or ART [8]. MSM engaged in HIV screening at regular intervals; we calibrated screening rates to reproduce 2017 estimates from the Georgia Department of Public Health (GDPH) surveillance report of the proportion of MSM with HIV who were diagnosed [31]. These calibrated screening rates were externally validated by comparing simulations to projections of national estimates of the median time between infection and diagnosis [32]. MSM screening positive could then enter the HIV care continuum (linkage and retention in ART care) while MSM who screened negative could enter the HIV prevention continuum (PrEP initiation, adherence, and persistence).

In the care continuum, MSM were linked to ART based on race-specific rates calibrated to GDPH surveillance data on proportions of MSM linked to care within 30 days of HIV diagnosis [31]. We assumed immediate initiation of ART at care linkage based on current HIV treatment guidelines [33]. After linkage, MSM could cycle off and back on ART; halting rates were calibrated to GDPH surveillance data levels of VL suppression [31]. ART reinitiation rates were kept fixed for reasons of model identifiability (reinitiation rates were negatively correlated with halting rates). MSM who remained on ART achieved suppression after 3 months of use [24]. Upon stopping ART, VL would rebound to set-point VL [34].

The HIV prevention continuum consisted of initiation, adherence, and persistence in PrEP care for daily oral TDF/FTC [4]. HIV-negative MSM who met indications for PrEP based on CDC guidelines were eligible to start [35]. Because MSM enter PrEP care through regular HIV screening or spontaneously [1], we modeled the link between HIV screening and PrEP in these two ways to understand the relationship between HIV screening and PrEP coverage. In the first scenario, MSM were eligible to start PrEP if they both had indications and they had screened negative for HIV that week. In the second scenario, MSM were eligible to start PrEP based on indications alone, so PrEP initiation was spontaneous and not linked to regular screening (however, confirmation of HIV-negative status was conducted upon PrEP initiation). In each scenario, eligible MSM then started PrEP based on an initiation probability generating an overall coverage level of 15%, consistent with estimates of Atlanta MSM [1].

After PrEP initiation, differential adherence was modeled, with 78.4% meeting the high-adherence level [36] that resulted in a 99% relative reduction in HIV acquisition risk [27]. MSM with high PrEP adherence reduced their condom use, based on ARTnet estimates (see Appendix Section 4.2). PrEP discontinuation rates were based on external estimates of the proportion of MSM who were retained in PrEP care at 6 months (57%) [37], transformed into median time to discontinuation (224 days). MSM additionally stopped PrEP if they no longer exhibited PrEP indications [35].

### Intervention Scenarios

In our primary scenarios, we varied the rates of HIV screening, ART care linkage, and ART care retention from the base race-stratified rates to simulate interventions for a 10-year period (2020 to 2030). For HIV screening, we modeled the two separate scenarios based on assumptions that PrEP initiation was linked to regular screening versus unlinked. For both scenarios, we simulated relative increases in current race-stratified screening rates as well as standardized target screening intervals (annual, biannual, quarterly) that were equal across race groups. We took a similar approach in the care linkage scenarios, while we only varied retention rates relative to base rates as there are no published target intervals. We also evaluated the impact of the same improvements in HIV care and prevention assuming those interventions were implement only among black MSM, the racial/ethnic subgroup with the highest local HIV burden [18].

We then considered the impact of combined improvements. First, we varied the relative screening and retention rates together to explore the combination of the two interventions, with two separate models varying the assumption about whether HIV screening and PrEP initiation are linked. Second, we simulated the most optimistic scenario considered, a 10-fold improvement in both screening and retention with PrEP linked to screening for a 50-year period to estimate how long it would take to reach the national EHE goal in the modeled population of MSM in Atlanta if these improvements were sustained.

### Simulation and Analysis

For each scenario, we simulated the model 1000 times and summarized the distribution of results with medians and 95% simulation intervals (SI). Primary outcomes were overall and race-stratified HIV incidence per 100 person-years at risk (PYAR) during the final intervention year. The model was based on three mutually exclusive race/ethnic groups, with proportions drawn from 2018 Census data for men living in the Atlanta metropolitan area: 51.5% non-Hispanic black, 4.6% Hispanic, and 43.9% non-Hispanic white/other. We also calculated the cumulative percent of infections averted (PIA) comparing the cumulative incidence over the intervention period for each counterfactual scenario to that in the base scenario. Process outcomes associated with each scenario included measures of the HIV care continuum (proportion of MSM with HIV who were screened, linked to care, and HIV virally suppressed at the end of the intervention scenario) and prevention continuum (current PrEP coverage).

## RESULTS

Table 1 presents the scenarios in which PrEP initiation was linked to HIV screening. Increases to the relative screening rates were associated with lower HIV incidence and higher PIA. A 10-fold increase in screening, which corresponds to increasing to approximately quarterly screening for white MSM and biannual screening for black and Hispanic MSM, resulted in a reduction in incidence from 1.19 to 0.66 per 100 PYAR overall, with 43.1% infections averted. The impact on incidence was non-linear across the relative rates, with the greatest difference in PIA at lower relative increases in rates: the difference in the PIA comparing base rates to 2-fold (14.7%) was larger than a doubling of rates from 5-fold to 10-fold (10.9%). By race/ethnicity, black MSM had the highest base HIV incidence (2.41 per 100 PYAR), followed by Hispanic MSM (0.61 per 100 PYAR), and white MSM (0.35 per 100 PYAR). With a 10-fold relative change to screening rates, a higher percent of infections would be averted for white MSM (53.5%) compared to Hispanic MSM (47.1%) and black MSM (40.8%) because of the larger absolute increase in screening for white MSM. Supplemental Table 14 shows the impact of PrEP-linked HIV screening increases if this intervention focused only on black MSM. Increasing HIV screening only among black MSM would avert a similar percentage of infections compared to the non-targeted screening of all MSM (36.8% versus 43.1%). This is due to the higher HIV prevalence among Black MSM, the relatively high proportion of the MSM population that is black, and the disassortative racial mixing in sexual partnership (so that prevention delivered to Black MSM also has indirect effects on non-Black MSM).

**Table 1.** HIV Incidence Associated with Changes to Relative Screening Rates and Average HIV Screening Interval, Overall and Stratified by Race/Ethnicity, with PrEP Initiation Linked to HIV Screening

| Scenario | Overall |  | Black MSM |  | Hispanic MSM |  | White MSM |  |
| --- | --- | --- | --- | --- | --- | --- | --- | --- |
|  | HIV Incidence | PIA<sup>B</sup> | HIV Incidence | PIA<sup>B</sup> | HIV Incidence | PIA<sup>B</sup> | HIV Incidence | PIA<sup>B</sup> |
|  | Rate<sup>A</sup> (95% SI) | % (95% SI) | Rate<sup>A</sup> (95% SI) | % (95% SI) | Rate<sup>A</sup> (95% SI) | % (95% SI) | Rate<sup>A</sup> (95% SI) | % (95% SI) |
| **Relative Rates** |  |  |  |  |  |  |  |  |
| Base Rates<sup>C</sup> | 1.19 (0.92, 1.48) | – | 2.41 (1.83, 3.05) | – | 0.61 (0.00, 1.60) | – | 0.35 (0.19, 0.54) | – |
| Base Rates x 2 | 0.99 (0.75, 1.25) | 14.7 (14.5, 15.0) | 1.99 (1.47, 2.60) | 13.8 (13.6, 14.1) | 0.58 (0.00, 1.38) | 16.2 (15.2, 17.1) | 0.27 (0.13, 0.44) | 18.8 (18.4, 19.2) |
| Base Rates x 5 | 0.78 (0.58, 1.01) | 32.2 (32.0, 32.4) | 1.59 (1.15, 2.11) | 30.3 (30.1, 30.5) | 0.39 (0.00, 1.14) | 35.3 (34.4, 36.1) | 0.19 (0.08, 0.34) | 41.1 (40.8, 41.5) |
| Base Rates x 10<sup>D</sup> | 0.66 (0.48, 0.85) | 43.1 (42.9, 43.2) | 1.34 (0.97, 1.77) | 40.8 (40.6, 41.0) | 0.38 (0.00, 0.99) | 47.1 (46.4, 47.6) | 0.15 (0.06, 0.27) | 53.5 (53.2, 53.8) |
| **Target Intervals** |  |  |  |  |  |  |  |  |
| Annual | 0.80 (0.59, 1.02) | 30.5 (30.4, 30.7) | 1.58 (1.17, 2.07) | 30.2 (30.0, 30.4) | 0.39 (0.00, 1.16) | 35.0 (34.2, 35.6) | 0.23 (0.11, 0.38) | 31.5 (31.1, 31.8) |
| Biannual | 0.67 (0.48, 0.88) | 42.0 (41.8, 42.2) | 1.32 (0.93, 1.79) | 41.2 (41.0, 41.3) | 0.38 (0.00, 0.98) | 46.7 (45.9, 47.2) | 0.17 (0.08, 0.32) | 45.1 (44.8, 45.4) |
| Quarterly | 0.56 (0.41, 0.73) | 50.9 (50.8, 51.1) | 1.11 (0.80, 1.50) | 49.8 (49.6, 49.9) | 0.20 (0.00, 0.83) | 56.1 (55.6, 56.8) | 0.15 (0.06, 0.26) | 55.4 (55.1, 55.6) |

Abbreviations: PIA, percent of infections averted; SI, simulation interval
<sup>A</sup> HIV incidence rate per 100 person-years at risk in the year 10 of the simulated intervention.
<sup>B</sup> Percent of infections averted over the 10-year intervention relative to the reference model.
<sup>C</sup> Base rates per week are 0.00385, 0.00380, and 0.00690 for black, Hispanic, and white MSM, corresponding to an average inter-test interval of 5.00, 5.06, and 2.79 years, respectively (see Supplemental Table 10).
<sup>D</sup> 10-fold rates per week are 0.0385, 0.0380, and 0.0690 for black, Hispanic, and white MSM, corresponding to an average inter-test interval of 26.0, 26.3, and 14.5 weeks, respectively.

The process mechanisms for incidence reduction here are shown in Supplemental Tables 15–18. HIV prevention occurred through a combined reduction in risks of HIV acquisition and transmission. Reduced HIV acquisition risk occurred through screening-driven increases in PrEP coverage, from 15.0% in the base scenario to 67.0% with a 10-fold increase in screening. As screening occurred more often, MSM had a greater opportunity to initiate PrEP during periods where they were tested and also met behavioral indications for PrEP. Standardizing screening rates and the offer of PrEP across race groups removed disparities in PrEP coverage. Reduced HIV transmission rates occurred through increased VL suppression that lowered the probability of transmission. Screening specifically increased the prevalence of suppression among all MSM (from 48.8% in the base scenario to 55.5% in the 10-fold scenario) given fixed linkage and retention rates. Annual screening increased the proportion diagnosed from 83.5% to 98.7%, and the median delay between infection and diagnosis decreased from 2.7 years to 0.7 years.

Table 2 shows the same set of screening rate counterfactuals under the assumption that PrEP initiation is not linked to HIV screening. In these scenarios a 10-fold increase in screening rates led to 10.9% of all infections being averted. This was one-quarter the prevention impact of the PrEP-linked scenario with the same relative increase in screening (10.9% versus 43.1% infections averted). By race/ethnicity, the reductions in incidence did not vary substantially for the relative rate counterfactuals. For the target screening intervals, annual screening averted 9.2% infections and quarterly screening averted 12.2% infections. Supplemental Table 19 shows the impact of PrEP-unlinked increases in HIV screening when focused only on black MSM; again, this strategy of focusing interventions to increase HIV screening on black MSM achieves nearly the same overall impact as non-targeted HIV screening.

**Table 2.** HIV Incidence Associated with Changes to Relative Screening Rates and Average HIV Screening Interval, Overall and Stratified by Race/Ethnicity, with Random PrEP Initiation (Not Linked to HIV Screening)

| Scenario | Overall |  | Black MSM |  | Hispanic MSM |  | White MSM |  |
| --- | --- | --- | --- | --- | --- | --- | --- | --- |
|  | HIV Incidence | PIA<sup>B</sup> | HIV Incidence | PIA<sup>B</sup> | HIV Incidence | PIA<sup>B</sup> | HIV Incidence | PIA<sup>B</sup> |
|  | Rate<sup>A</sup> (95% SI) | % (95% SI) | Rate<sup>A</sup> (95% SI) | % (95% SI) | Rate<sup>A</sup> (95% SI) | % (95% SI) | Rate<sup>A</sup> (95% SI) | % (95% SI) |
| **Relative Rates** |  |  |  |  |  |  |  |  |
| Base Rates<sup>C</sup> | 1.09 (0.84, 1.40) | – | 2.17 (1.60, 2.81) | – | 0.58 (0.00, 1.37) | – | 0.32 (0.18, 0.49) | – |
| Base Rates x 2 | 1.03 (0.80, 1.30) | 5.0 (4.7, 5.2) | 2.03 (1.54, 2.60) | 5.0 (4.7, 5.3) | 0.57 (0.00, 1.37) | 5.9 (3.8, 6.7) | 0.30 (0.16, 0.50) | 4.8 (4.3, 5.3) |
| Base Rates x 5 | 1.00 (0.76, 1.26) | 9.3 (9.1, 9.6) | 1.96 (1.45, 2.55) | 9.3 (9.1, 9.6) | 0.57 (0.00, 1.33) | 10.7 (9.7, 11.8) | 0.29 (0.14, 0.47) | 9.5 (9.1, 10.1) |
| Base Rates x 10<sup>D</sup> | 0.99 (0.75, 1.27) | 10.9 (10.6, 11.1) | 1.95 (1.44, 2.52) | 10.8 (10.5, 11.0) | 0.56 (0.00, 1.20) | 12.5 (11.4, 13.5) | 0.28 (0.14, 0.45) | 11.0 (10.6, 11.5) |
| **Target Intervals** |  |  |  |  |  |  |  |  |
| Annual | 1.00 (0.75, 1.27) | 9.2 (8.9, 9.4) | 1.96 (1.43, 2.56) | 9.2 (8.9, 9.4) | 0.57 (0.00, 1.34) | 10.7 (9.7, 11.8) | 0.29 (0.14, 0.46) | 9.1 (8.6, 9.5) |
| Biannual | 0.99 (0.73, 1.24) | 11.2 (11.0, 11.4) | 1.93 (1.41, 2.47) | 11.1 (10.9, 11.4) | 0.57 (0.00, 1.36) | 12.1 (11.0, 13.0) | 0.29 (0.14, 0.47) | 11.6 (11.1, 12.1) |
| Quarterly | 0.99 (0.76, 1.26) | 12.2 (11.9, 12.4) | 1.94 (1.47, 2.51) | 11.9 (11.7, 12.2) | 0.57 (0.00, 1.31) | 13.2 (12.1, 14.3) | 0.29 (0.14, 0.47) | 13.1 (12.6, 13.6) |

Abbreviations: PIA, percent of infections averted; SI, simulation interval
<sup>A</sup> HIV incidence rate per 100 person-years at risk in the year 10 of the simulated intervention.
<sup>B</sup> Percent of infections averted over the 10-year intervention relative to the reference model.
<sup>C</sup> Base rates per week are 0.00385, 0.00380, and 0.00690 for black, Hispanic, and white MSM, corresponding to an average inter-test interval of 5.00, 5.06, and 2.79 years, respectively (see Supplemental Table 10).
<sup>D</sup> 10-fold rates per week are 0.0385, 0.0380, and 0.0690 for black, Hispanic, and white MSM, corresponding to an average inter-test interval of 26.0, 26.3, and 14.5 weeks, respectively.

The process mechanisms shown in Supplemental Tables 20–23 demonstrate that, unlike in the PrEP-linked scenarios, PrEP coverage did not vary with increases to HIV screening: it remained constant at ∼15% overall. Here the only prevention mechanisms are through reduction in transmission risk through changes to VL suppression levels, from 51.0% in the base scenario to 56.5% in the 10-fold scenario. Because these changes in VL suppression were similar to those in the PrEP-linked scenarios, the difference in infections averted between the two scenario sets was attributable to increases in screening-driven PrEP coverage.

Table 3 presents the model scenarios for HIV care linkage and retention. Compared to base rates of HIV care linkage within 30 days of diagnosis, a 2-fold increase in rates had little impact on HIV incidence. Even immediate linkage averted fewer than 1% of infections compared to current estimated linkage rates. The impact of care linkage interventions did not vary by race/ethnicity. Supplemental Table 24 shows the intervention targeted at Black MSM. Mechanisms are shown in Supplemental Tables 25–28. Although doubling the rate of care linkage would increase the proportion linked to care within one month of diagnosis from 64.8% to 90.1%, this by itself (i.e., with HIV care retention fixed) had no impact on the overall levels of VL suppression.

**Table 3.** HIV Incidence Associated with Changes to HIV Care Linkage Rates and HIV Care Retention Rates, Overall and Stratified by Race/Ethnicity

| Scenario | Overall |  | Black MSM |  | Hispanic MSM |  | White MSM |  |
| --- | --- | --- | --- | --- | --- | --- | --- | --- |
|  | HIV Incidence | PIA<sup>B</sup> | HIV Incidence | PIA<sup>B</sup> | HIV Incidence | PIA<sup>B</sup> | HIV Incidence | PIA<sup>B</sup> |
|  | Rate<sup>A</sup> (95% SI) | % (95% SI) | Rate<sup>A</sup> (95% SI) | % (95% SI) | Rate<sup>A</sup> (95% SI) | % (95% SI) | Rate<sup>A</sup> (95% SI) | % (95% SI) |
| **HIV Care Linkage** |  |  |  |  |  |  |  |  |
| *Relative Rates* |  |  |  |  |  |  |  |  |
| Base Rates<sup>C</sup> | 1.18 (0.91, 1.48) | – | 2.40 (1.80, 3.09) | – | 0.60 (0.00, 1.52) | – | 0.35 (0.21, 0.54) | – |
| Base Rate x 1.5 | 1.19 (0.92, 1.52) | 0.0 (-0.3, 0.3) | 2.40 (1.79, 3.15) | -0.1 (-0.4, 0.2) | 0.61 (0.00, 1.60) | 0.0 (-2.5, 0.0) | 0.35 (0.19, 0.56) | 0.5 (0.0, 1.0) |
| Base Rate x 2 | 1.19 (0.92, 1.50) | -0.1 (-0.4, 0.2) | 2.40 (1.82, 3.03) | -0.1 (-0.4, 0.1) | 0.61 (0.00, 1.46) | 0.0 (-2.6, 0.0) | 0.35 (0.19, 0.54) | 0.0 (-0.5, 0.5) |
| *Interval Targets* |  |  |  |  |  |  |  |  |
| One Month | 1.18 (0.92, 1.47) | 0.1 (-0.2, 0.4) | 2.39 (1.80, 3.04) | 0.1 (-0.2, 0.4) | 0.61 (0.19, 1.55) | 0.0 (-2.5, 0.0) | 0.34 (0.19, 0.54) | 0.0 (-0.5, 0.6) |
| Two Weeks | 1.19 (0.91, 1.47) | 0.2 (-0.1, 0.4) | 2.37 (1.82, 3.03) | 0.1 (-0.1, 0.4) | 0.61 (0.00, 1.58) | 0.0 (0.0, 0.0) | 0.35 (0.19, 0.52) | 0.5 (0.0, 1.0) |
| Immediate | 1.17 (0.92, 1.46) | 0.8 (0.5, 1.0) | 2.36 (1.79, 3.05) | 0.7 (0.4, 1.0) | 0.61 (0.00, 1.57) | 0.0 (0.0, 2.3) | 0.34 (0.19, 0.53) | 1.0 (0.5, 1.5) |
| **HIV Care Retention** |  |  |  |  |  |  |  |  |
| *Relative Rates* |  |  |  |  |  |  |  |  |
| Base Rates<sup>D</sup> | 1.18 (0.90, 1.50) | – | 2.39 (1.77, 3.07) | – | 0.61 (0.00, 1.57) | – | 0.35 (0.19, 0.56) | – |
| Base Rates x 1.5 | 0.97 (0.73, 1.25) | 12.1 (11.8, 12.3) | 1.95 (1.43, 2.54) | 11.8 (11.6, 12.1) | 0.57 (0.00, 1.27) | 13.9 (13.0, 14.9) | 0.29 (0.14, 0.46) | 12.9 (12.4, 13.4) |
| Base Rates x 2 | 0.84 (0.62, 1.09) | 19.1 (18.9, 19.4) | 1.67 (1.20, 2.22) | 19.0 (18.7, 19.2) | 0.40 (0.00, 1.19) | 20.6 (19.8, 21.6) | 0.23 (0.11, 0.38) | 19.8 (19.3, 20.2) |
| Base Rates x 4 | 0.60 (0.42, 0.83) | 31.5 (31.2, 31.7) | 1.19 (0.78, 1.69) | 31.3 (31.1, 31.6) | 0.38 (0.00, 0.98) | 32.5 (32.1, 33.3) | 0.17 (0.06, 0.31) | 31.9 (31.5, 32.3) |
| Base Rates x 10<sup>E</sup> | 0.42 (0.26, 0.61) | 40.5 (40.3, 40.7) | 0.82 (0.49, 1.22) | 40.4 (40.2, 40.6) | 0.20 (0.00, 0.79) | 41.6 (40.7, 42.3) | 0.11 (0.02, 0.23) | 40.6 (40.2, 41.0) |

Abbreviations: PIA, percent of infections averted; SI, simulation interval
<sup>A</sup> HIV incidence rate per 100 person-years at risk in the year 10 of the simulated intervention.<sup>B</sup> Percent of infections averted over the 10-year intervention relative to the reference model. PIA may be negative if the cumulative incidence in the experimental scenario is higher than the cumulative incidence in the reference scenario.<sup>C</sup> Base rates per week are 0.1775, 0.1900, and 0.2521 for black, Hispanic, and white MSM, corresponding to an average time to linkage of 5.6, 5.3, and 4.0 weeks, respectively (see Supplemental Table 11).<sup>D</sup> Base rates per week are 0.0062, 0.0055, and 0.0031 for black, Hispanic, and white MSM, corresponding to an average time on ART before stoppage of 3.1, 3.5, and 6.2 years, respectively.<sup>E</sup> 10-fold rates per week are 0.062, 0.055, and 0.031 for black, Hispanic, and white MSM, corresponding to an average time on ART before stoppage of 31, 35, and 62 years, respectively.

Alternatively, increases in care retention, resulted in substantial reductions in HIV incidence (Table 3). Overall, a 2-fold increase in retention rates would avert 19.1% of infections and a 10-fold increase would avert 40.5% of infections. The time to first ART stoppage in the reference scenario was 3.1 years, 3.5 years, and 6.2 years for black, Hispanic, and white MSM (see Supplemental Table 12), so a 10-fold increase would represent remaining on ART for 31, 35, and 62 years on average, respectively. By race/ethnicity, there were only minor differences in the PIAs across the rate scenarios, with 40.4% to 41.6% averted in the 10-fold improvement scenario. Focusing care retention interventions on black MSM would lead to similar overall prevention benefits to an intervention that improved retention for all MSM (Supplemental Table 24).

The mechanisms for retention on incidence reduction are shown in Supplemental Tables 29–32. Increases in retention result in large increases in the fraction of HIV-diagnosed MSM and all MSM with HIV with VL suppression: a 10-fold rate increase would result in 89.2% of diagnosed MSM achieving VL suppression. As the race/ethnicity-stratified tables show, although there were large differences in VL suppression in the base scenarios, a 10-fold increase in the retention rates led to nearly 90% of those with diagnosed infection in each race/ethnicity group achieving viral suppression.

Figure 1 visualizes the PIA when screening rates and retention rates were jointly varied up to 10-fold from current estimates. The two panels contrast the effects under assumptions of PrEP-linked screening. If both screening and retention were increased 10-fold, 73.9% and 57.9% of infections were averted under assumptions of PrEP linkage and no-linkage, respectively. In the PrEP-linked scenario, the vertical contours at lower levels of screening demonstrate the greater relative benefit of increased screening (with PrEP initiation). In the PrEP-unlinked scenario, the horizontal contours show the relatively limited benefit of increased screening (without PrEP increases) compared to HIV care retention. Numerical results are provided in Supplemental Table 33. In the model with model screening and retention rates increased 10-fold, the proportion of MSM with HIV who were diagnosed exceeded 99% and the proportion of the diagnosed who were virally suppressed was 89%.

**Figure 1.**
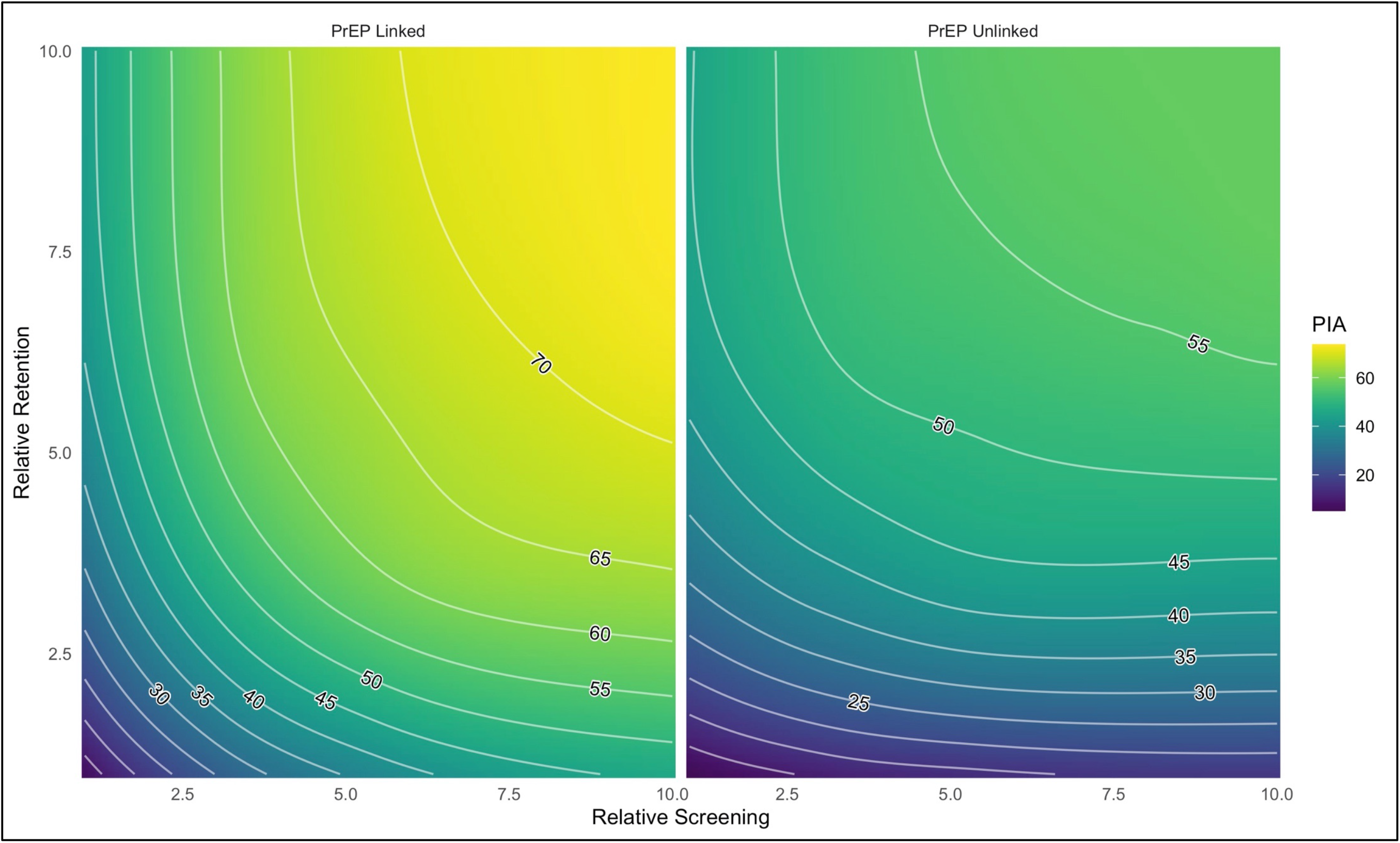
Percent of cumulative infections averted (PIA) over 10 years as a function of relative improvements to HIV screening rates and HIV care retention rates, under a scenario where the offer of PrEP is linked to screening negative (left panel) or that the offer of PrEP occurs as a separate activity with no linkage to HIV screening (right panel). The maximum PIA achieved (upper right corners of each panel) were 73.9% and 57.9%, respectively.

Because only a 74% cumulative incidence reduction was achieved after 10 years in the most optimistic scenario in Figure 1 (the upper right corner of the left panel), Figure 2 displays simulating this scenario out for 50 years to evaluate how long it would take to reach the EHE 2025 75% reduction goal and the 2030 90% reduction goal in point incidence. The reference model held the HIV prevention and care continuum levels (constant whereas the 10×10 model increased HIV screening (with linked PrEP) and retention rates 10-fold. We projected that 75% incidence reduction goal would be achieved in year 2024 (4.0 years post-intervention initiation) and the 90% incidence reduction goal would be achieved in 2032 (12.0 years post-intervention initiation).

**Figure 2.**
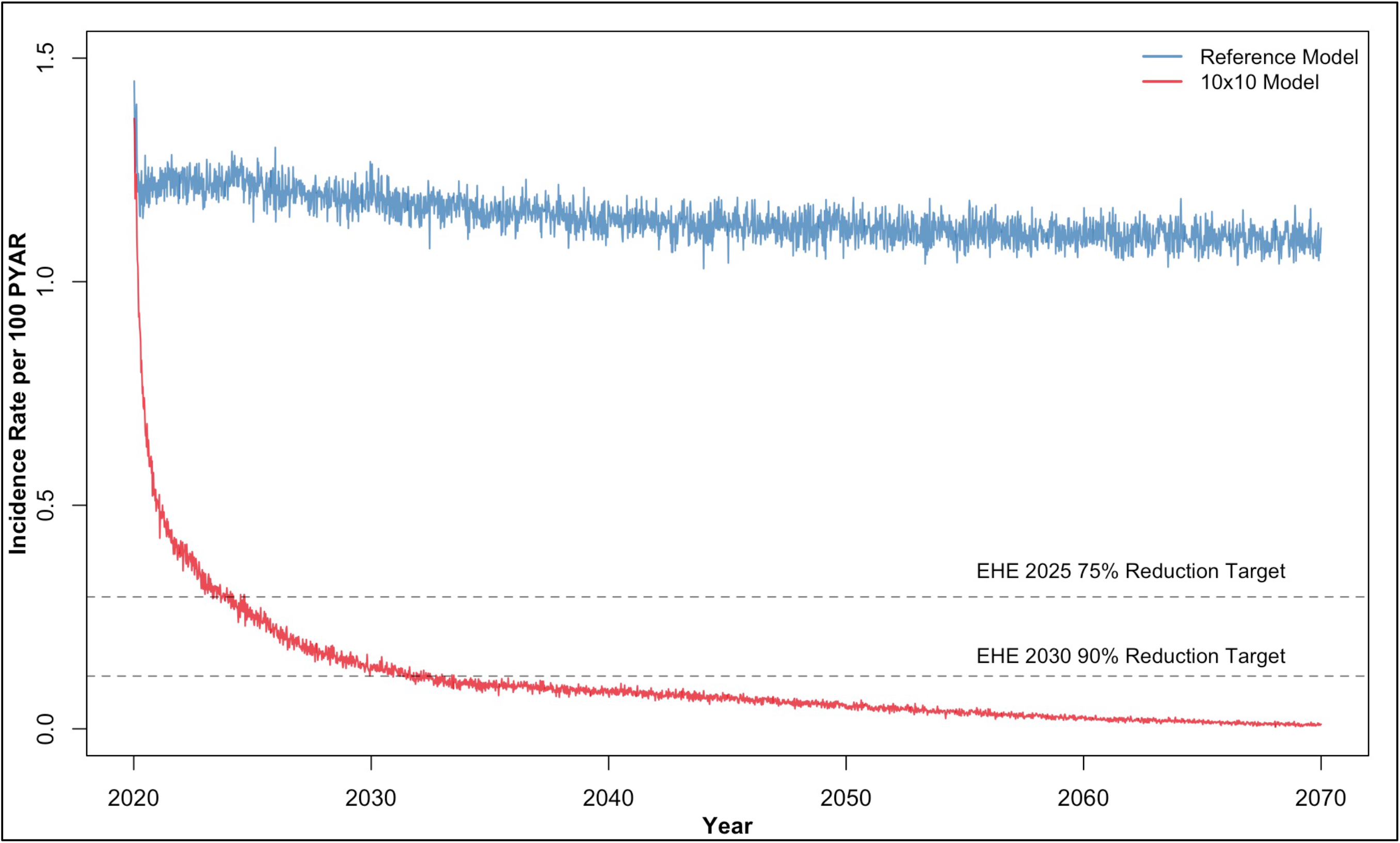
HIV incidence rate per 100 person-years at risk (PYAR) over 50 years comparing a scenario in which the HIV prevention and care continuum was fixed (Reference Model) to a scenario in which there were 10-fold increases in rates of HIV screening (with linked PrEP) and care retention (10×10 Model). The EHE target of a 75% reduction in the incidence rate is achieved in the 10×10 Model in year 2027 (4 years post-intervention initiation) and the 90% incidence reduction target is achieved in 2032 (12 years post-intervention initiation).

## DISCUSSION

In this study, we found that major improvements to an integrated HIV prevention and care continuum could avert over two-thirds of HIV infections expected among MSM in Atlanta over the next decade. HIV screening interventions combined with HIV PrEP linkage would provide a major prevention benefit compared to HIV screening not tied to PrEP. Although it may be possible to reach the 2025 75% incidence reduction goal by the 2025 target with this combination of large improvements in both the treatment and prevention continua, reaching the 90% was not projected to be possible by the 2030 target. Achieving the EHE goal in high-need areas like Atlanta will require a substantial acceleration of HIV prevention and care service delivery.

Several mathematical modeling studies have evaluated how meeting HIV care continuum targets in the United States impacts HIV incidence [38,39], while others have evaluated the effects of specific HIV prevention tools [14,15], but few modeling studies have evaluated packages of interventions and their synergistic effects on an integrated HIV prevention and care continuum. Previous care continuum models for the U.S. have generally found that improving HIV care retention would be higher impact than focusing on HIV screening or linkage [10,40,41], although that has not been universal [11,42]. A recent economic model estimated the cost-effectiveness of scaling-up interventions such as HIV screening, PrEP, and ART retention under the EHE initiative [17,43]. However, this model found weak impact of HIV PrEP potentially due to limitations of its compartmental framework and assumptions that each EHE activity was mechanistically unrelated to the others.

In contrast, our model investigated how the relative prevention benefits of screening versus retention depended upon separate versus integrated HIV prevention and care continua that specifically reflect these mechanistic interactions between these interventions. PrEP-linked screening, by lowering both acquisition and transmission risks, resulted in an impact of screening comparable to that of improving rates of retention for MSM with HIV. Previous models of HIV screening and linkage interventions for MSM with HIV have only included the reduction of HIV transmission risk through accelerated suppression of HIV VL [10]. The transmission risk reduction benefits of HIV screening and linkage interventions compared to HIV care retention interventions are transient: the median time between infection and diagnosis, and between diagnosis and care linkage are approximately 2.5 years and 3 months [32]. HIV retention interventions may now span decades under appropriate ART [44].

However, even ambitious levels of screening alone may be insufficient to meet the EHE incidence reduction goals in this population. The current EHE plan has intermediate goals of 95% of persons living with HIV to be diagnosed and 95% of the diagnosed to be virally suppressed [5]. We found at least a 5-fold increase in current screening rates would be needed to achieve the former goal, while a 10-fold retention increase only yielded 89% for the viral suppression goal in this population. Our joint 10×10 scenario (Figure 2) suggests that combined of improvements to HIV screening linked with PrEP initiation and HIV care retention could yield major reductions in HIV incidence within a decade, but with slower improvements after that. Challenges in meeting the 2030 90% EHE incidence reduction goal may also reflect that our model was parameterized to the HIV epidemic for MSM in Atlanta, with high HIV incidence and low rates of PrEP and VL suppression at baseline. This may also suggest that more targeted approaches to HIV screening linked with PrEP and ART are also needed, including those detecting emerging HIV clusters [45].

### Limitations

First, current HIV screening practice is more heterogenous than the form represented in the model. Whereas we simulated race/ethnicity-stratified rates of interval-based screening, real-world HIV screening events occur through regular clinical check-ups and risk events (such as acquiring a new partner) [46]. Additionally, HIV surveillance data suggests a bimodal distribution of screening, where some MSM get routine screening and some MSM remain undiagnosed until AIDS [31,47]. We did not model this heterogeneity because of the complexity in representing the interaction of testing typologies with HIV progression rates; it would also complicate the design of the model scenarios. Second, we did not investigate the cost-effectiveness of the modeled interventions. Despite the epidemiological advantage of HIV screening linked with PrEP, the cost of medication would be considerable if PrEP coverage increased as we projected [48]. However, economic analyses for PrEP may be in a state of flux because of the unknown pricing of generic TDF/FTC as it becomes available in 2020, as well as new programs that incentivize PrEP access as part of the EHE initiative [49]. Third, our target population (MSM in the Atlanta area) was selected based on the availability of data for model parameterization, the burden of HIV, and the focus of the EHE in this jurisdiction. Some of the broader conclusions about the impact of interventions focused specifically on black MSM, and for combinations of interventions, may differ in other populations and may not be generalizable to the feasibility of achieving the EHE goals nationally.

### Conclusions

The Ending the HIV Epidemic strategy of first targeting areas with the highest number of new HIV diagnoses means that EHE will also need to be implemented in places with many challenges and existing missed opportunities for detection, prevention, and control of HIV [5]. EHE’s ambitious 2030 HIV prevention goals require substantial increases in access to and use of HIV diagnostic, prevention, and treatment tools.

## Data Availability

Model data and analysis scripts are available on our Github repository linked below.

https://github.com/EpiModel/CombPrev

## Disclaimer

The findings and conclusions in this manuscript are those of the authors and do not necessarily represent the official positions of the Centers for Disease Control and Prevention.

## Contributions

SMJ designed the study, conducted the analyses, and drafted the manuscript. JAJ, KWH, DKS, and KPH provided key input in model parameterization and structure, and critically edited the manuscript.

